# Concussion, attention, and mental health: A brain network study in youth (CAMBY) – Study protocol

**DOI:** 10.64898/2026.10.14.26365111

**Authors:** Shannon E. Scratch, Pavreet K. Gill, Andrea Hickling, Alexa Irvin, Rouzbeh Zamyadi, Shireen Parimoo, Phillip R. Johnston, Adrienne L. Davis, George M. Ibrahim, Benjamin T. Dunkley, Anne L. Wheeler

## Abstract

**BACKGROUND:** One in three adolescents with a concussion history report experiencing long-lasting somatic, sleep, emotional, and/or cognitive disturbances (persisting symptoms after concussion (PSaC)). Cognitive difficulties, such as attentional problems, are common symptoms and can interfere with ongoing development. Clinically, psychiatric symptoms also present in youth with PSaC. A concussion causes diffuse functional and microstructural alterations to neural networks. To develop targeted treatments for PSaC (e.g., neurostimulation), we need to understand how and where structural alterations and neural dysfunction intersect to cause long-lasting symptoms.

**OBJECTIVES:** 1) Identify functional and microstructural properties in intrinsic connectivity networks, as well as neuropsychological outcomes, that differentiate adolescents with concussion with PSaC from those who do not have PSaC. 2) Determine whether neural activity within intrinsic connectivity networks mediates the relationship between network microstructure and specific neuropsychological features in adolescents with concussion.

**METHODS:** CAMBY is a prospective study of adolescents aged 12 to 18 years with a history of concussion. Participants are allocated based on recovery status (PSaC vs. No-PSaC (recovered controls)). Advanced structural (magnetic resonance imaging; MRI) and functional (magnetoencephalography; MEG) neuroimaging and neuropsychological measures will be collected one month or more after concussion. Microstructural and functional features will be computed within intrinsic connectivity networks, then compared between PSaC and recovered control groups and related to symptom domains and neuropsychological profiles.

**IMPACT:** This will be the first study to integrate MRI, MEG, and neuropsychology in adolescents with PSaC to identify disrupted networks underlying PSaC. This observational study is a necessary step towards future clinical trials that accurately target promising therapeutic neurostimulation protocols.

## Introduction

Concussion in adolescents is a serious public health concern. A concussion is a mild traumatic brain injury caused by mechanical forces transferred to the brain from the head, neck or body, resulting in disruptions in brain function (1,2). In Canada, 1.6% of Canadians aged 12 years or older reported sustaining one or more concussions in 2019, with the highest incidence (5.6%) among those aged 12-19 years (3). The average rate of concussion-related hospitalizations in Ontario, Canada in 2024 was estimated to be 4.28 per 100,000, with 28.3% being sport-related (4). It is estimated that thirty percent of adolescents with concussion will experience persisting symptoms after concussion (PSaC), which extend beyond 4 weeks post injury (1,5). Adolescents are more likely to experience PSaC than children (6) or adults (7,8) and PSaC significantly lower health-related quality of life (7,9–11).

Youth diagnosed with PSaC commonly experience a combination of somatic, sleep, psychiatric and cognitive symptoms (12–16). Somatic symptoms (including headaches, dizziness, nausea, light or noise sensitivity) are common and may reflect disruptions in autonomic regulation, vestibular function, or sensory processing (17–20). Sleep disturbances can involve sleeping too much or too little which can exacerbate other symptom domains (21,22). Youth with PSaC present with many undiagnosed psychiatric symptoms, especially high levels of anxiety and low mood (14,15,23–25). Cognitive disruptions associated with PSaC can interfere with ongoing development (25)(26) by reducing a youth’s ability to acquire knowledge and skills, ultimately affecting educational and employment options (27–29). Attentional difficulties are among the most commonly reported cognitive problems in youth with a history of concussion (30,31). Because attention is a foundational process that supports other cognitive functions, such as memory and executive functioning (32–35), treating attentional impairments in adolescents with PSaC may have broad, downstream benefits.

Current management of PSaC involves multidisciplinary care targeting affected symptom domains (36); however, there are no brain-based therapeutic interventions that target PSaC (37,38). A handful of Repetitive Transcranial Magnetic Stimulation (rTMS) trials have yielded mixed results in PSaC (39–42), underscoring the need for mechanistically guided network targets.

Although the location of functional disruption after concussion varies widely across individuals, converging evidence shows that PSaC reflect dysfunction in large-scale neural networks rather than focal injury (43). Altered functional connectivity within intrinsic connectivity networks, including the salience, default mode, executive control, and somatomotor networks has been linked to symptom severity, impaired cognitive control, and emotional dysregulation (44,45). Though the majority of functional imaging studies in concussion use functional MRI (fMRI), cryogenic magnetoencephalography (MEG) provides a complementary window into neural circuit dynamics, and is sensitive to fast, millisecond-scale changes in membrane potentials and synaptic integration, processes not accessible with hemodynamic methods (46–49). MEG has revealed substantial neurophysiological abnormalities after concussion in both young adults and children (46–48,50–55).

Given its structural characteristics, white matter is highly vulnerable to primary and secondary injury processes initiated by concussion. Diffusion MRI is well suited to detect the subtle microstructural alterations in white matter and shows measurable differences in youth that relate to symptom burden (56–60). The localization of imaging evidence of microstructural disruption is similarly heterogeneous to the functional literature (61) however, recent evidence suggests that superficial white matter may have enhanced vulnerability in development due to its localization at the grey-white matter interface and protracted maturation (58,59,62).

PSaC are associated with network-level functional disturbances and microstructural alterations, but it remains unclear how these relate to each other to inform mechanistic interventions. Few existing studies integrate both structural and functional measures to clarify how microstructural changes affect network dynamics and distinct symptom domains, with many using uni-modal approaches (e.g., only MRI or only MEG), lacking neuropsychological assessments of cognition and mental health, or assessing only adult populations with small sample sizes (63–70). The combined use of MRI and MEG provides superior sensitivity and specificity for detecting the structural and functional disturbances that sustain PSaC and no existing pediatric study has applied this multimodal approach or linked network-level abnormalities to clinically meaningful symptom and neuropsychological profiles. This research fills a critical gap in the field and will establish the mechanistic foundation necessary for developing targeted, brain-based treatments for PSaC, including future network-guided neuromodulation trials.

### Objectives

By comparing network-based microstructure and neuronal function, as well as neuropsychological outcomes, in adolescents with concussion who do and do not have PSaC, we will

1. identify functional and microstructural properties in intrinsic connectivity networks that differentiate adolescents with PSaC from those who do not have PSaC (recovered controls)
2. determine whether neural activity within intrinsic connectivity networks mediates the relationship between network microstructure and distinct symptoms and neuropsychological features in adolescents with concussion.

## Methods

### Study Overview

This study uses a prospective cohort design of adolescents who sustained a concussion and (i) have not recovered and are experiencing persisting symptoms (PSaC group) and (ii) have recovered at the point of study participation (No-PSaC; Recovered Control group). One hundred and sixty adolescents, aged 12 to 18 years (18 years, 11 months), will be recruited from Holland Bloorview Kids Rehabilitation Hospital (HBKRH) and The Hospital for Sick Children (SickKids). Both groups will undergo the same study procedures that involve completing a battery of cognitive measures and psychological questionnaires (neuropsychological assessment), an MRI scan, and MEG imaging a minimum of four weeks after the concussion (Figure 1). Caregivers/parents of youth participants will complete parent-reported versions of questionnaires completed by the adolescent participants. Recruitment began in April 2024 and will occur for approximately 3 years.

**Figure 1.**
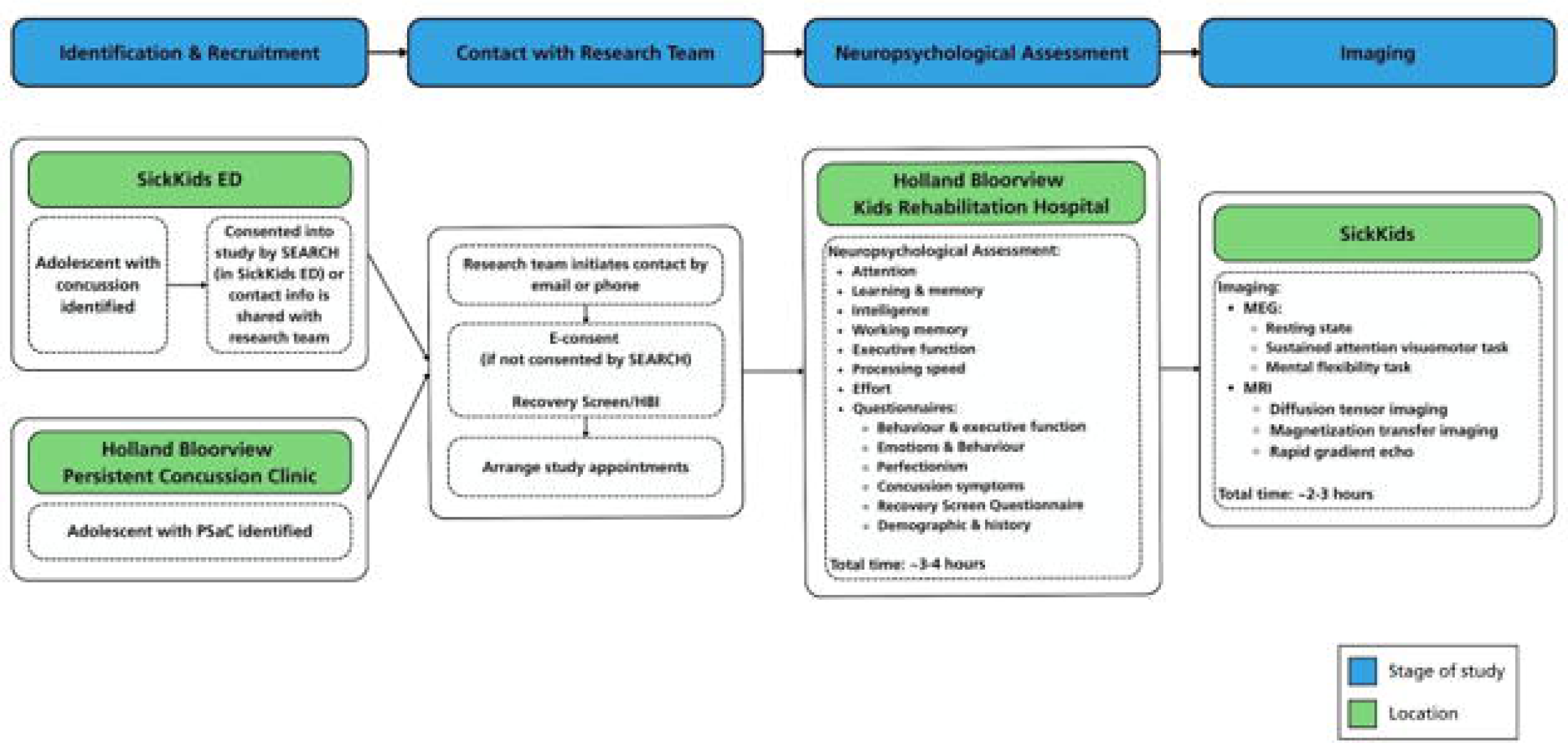
Schematic of participant recruitment and data collection. Participants are recruited from The Hospital for Sick Children (SickKids) and Holland Bloorview Kids Rehabilitation Hospital. Participants participate in a neuropsychological assessment and brain imaging a minimum of four weeks after their concussion.

### Participants

#### Inclusion Criteria

Eligible adolescents are 12-18 years old and have been diagnosed with a concussion by a physician or nurse practitioner at any point before participating. Eligible caregivers are the caregiver of a youth who participates in the study and have the capacity to consent and complete the requested questionnaires.

Based on self-report, participants will be assigned to the PSaC group if they: 1) have not returned to their pre-concussion levels of participation in school, work, and/or extracurriculars (including sport), and 2) endorse a high symptom burden, defined as 3+ symptoms at the time of study participation, accounting for normal symptom variation that can be observed in daily life (71,72).

Based on self-report, participants will be assigned to the recovered control group if they have self-reportedly returned to pre-concussion levels of school, work, and extracurriculars (including sport) at the time of their study participation.

In line with clinical diagnostic criteria (1,5,36,73), we will take a multi-pronged and conservative approach when determining if participants have PSaC and evaluate participants in two ways: (1) functionally (i.e., return or lack of return to activity), and (2) symptomatically (i.e., self-reported ratings of symptom burden). Analyses will be carried out with these two approaches in mind.

We acknowledge that there is variation in this population due to the complexity of concussion presentation. Thus, a partially recovered group may present once all data has been collected. If partial recovery is identified, sensitivity analyses will be conducted.

#### Exclusion Criteria

Exclusion criteria for all participants include standard MRI contraindications (74).

Contraindications include implantable medical devices (e.g., cardiac pacemaker, hearing aid), metallic fragments within the body, braces and some dental work/wires that can cause image distortion. Participants will also be excluded if they are unable to read or speak English, are unable to provide assent/consent, or have been diagnosed with a co-morbid developmental condition that would impair an individual’s ability to participate in neuropsychological testing (e.g. intellectual disability, global developmental delay, severe communication disorder, autism spectrum disorder with severe language impairment, in-patients receiving active medical/mental health treatment). Participants will also be excluded if they have ever experienced a moderate or severe brain injury or had a positive actionable finding during previous research or clinical brain scans.

#### Recruitment

Participants will be primarily recruited from Holland Bloorview Kids Rehabilitation Hospital (HBKRH) and SickKids in Toronto, Ontario, Canada. At HBKRH, adolescents will be recruited through Concussion Services, which provides multidisciplinary services to children, adolescents, and young adults experiencing PSaC (25,75,76). Recruitment will also take place through HBKRH’s Connect2Research database. Connect2Research is a secure database that contains limited information, including date of birth, diagnosis, and contact information of past and present clients and families of HBKRH. At SickKids, participants will be recruited in the Emergency Department by the SickKids Emergency Assistants for Research in Child Health (SEARCH) Program. After confirming eligibility with the clinical team, SEARCH will either obtain consent from the patient in the emergency department, schedule an onboarding call for the eligible patient with the research team, or obtain consent to share the eligible patient’s contact information with the research team for follow-up. Participants may also be recruited through local community clinics/organizations or snowball recruitment. Recruitment started on April 1st, 2024 and is expected to end March 31st, 2027.

### Consent

As youth and caregivers will undergo separate data collection procedures, separate consent procedures will be used for each participant group. Youth and caregiver participants will provide written consent following an informed discussion. In Ontario, Canada, there is no age of consent. For youth who do not demonstrate capacity to consent, verbal and written assent will be obtained with additional written consent obtained from a substitute decision maker (i.e., parent or caregiver). All consent forms will be signed and stored electronically via REDCap. Participants will also be invited to sign an optional consent form that allows the research team to share their de-identified study data to the Health Data Nexus database (77) for future unspecified research. Health Data Nexus is managed by the Temerty Centre for AI Research and Education in Medicine (T-CAIREM) at the University of Toronto (https://healthdatanexus.ai/).

### Data Collection Procedures

#### Overview

All youth participants (PSaC group and recovered control group) will undergo the same data collection procedures a minimum of four weeks post-concussion: 1) a neuropsychological assessment at HBKRH covering a range of cognitive domains and questionnaires to characterize symptoms and mental health, and 2) MRI and MEG imaging at SickKids (Figure 1). Assessments will occur a maximum of two weeks apart. A caregiver of each youth participant will also complete questionnaires. Data collection is currently ongoing and expected to be completed by March 2027 with completion of recruitment.

#### Symptom assessments

Concussion symptoms and concussion-related functioning will be captured via the Health and Behaviour Inventory (HBI) (78,79) and the study-developed Recovery Screen, respectively. The HBI and Recovery Screen will be administered at multiple timepoints to capture changes in self-reported symptoms and functioning post-concussion (Figure 2). Three versions of the youth and parent-report HBI will be used: (1) *the retrospective version* captures concussion symptoms in the week prior to injury, (2) *the acute version* captures concussion symptoms since the time of injury, and (3) *the follow-up version* captures concussion symptoms in the past week. Youth (and their caregivers) who are enrolled in the study less than four weeks (≤28 days) post-concussion will fill out the *retrospective and acute version* of the HBI upon consenting into the study, and then complete the *follow-up version* five weeks post-concussion and/or at the time of the neuropsychological assessment. Youth (and their caregivers) who enroll in the study more than four weeks (>28 days) post-concussion fill out the *follow-up version* upon consenting and also complete the *follow-up version* again at the neuropsychological assessment. The retrospective version was not completed past 28 days as retrospective reliability decreases as post-injury time increases (80–82).

**Figure 2.**
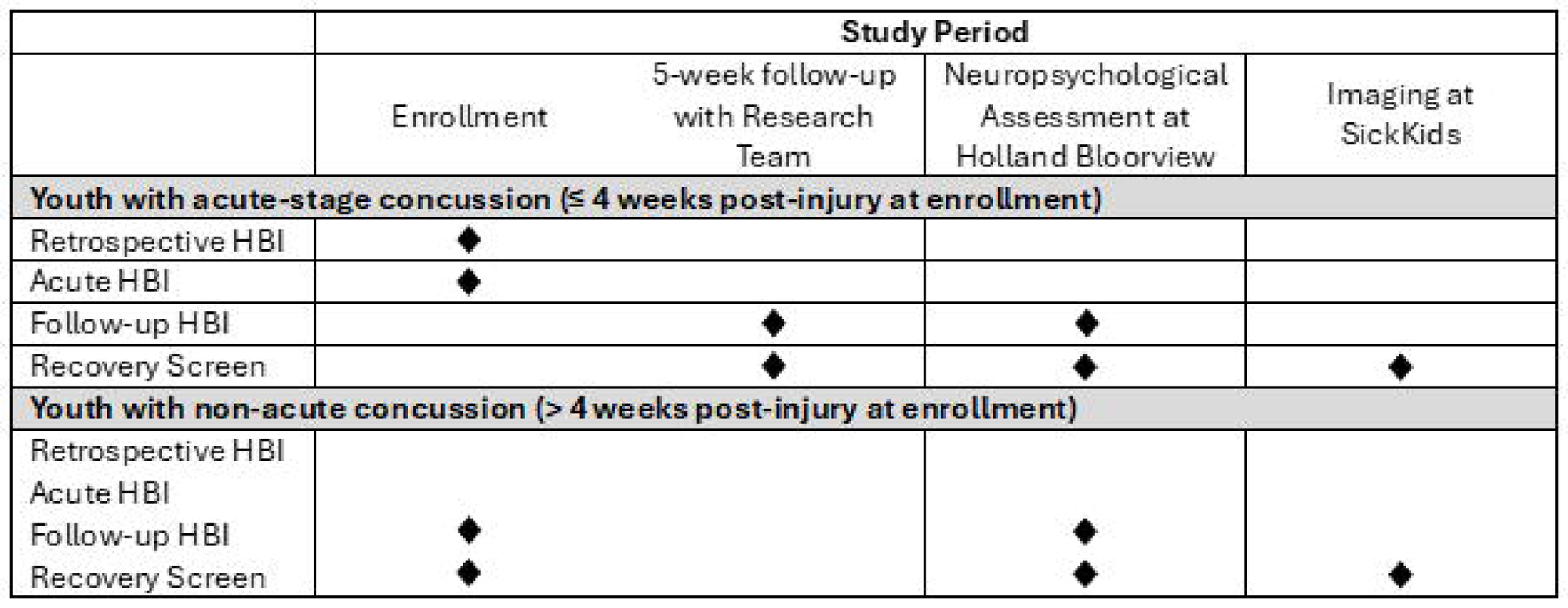
Administration of the Health and Behaviour Inventory (HBI) and Recovery Screen for youth participants. Each time a youth fills out the HBI, the parent/caregiver participant will also complete the accompanying parent version of the form.

The Recovery Screen is a three item (yes/no) questionnaire designed by the research team that aims to assess if a youth has regained self-reported typical or pre-injury levels of function at school, work, and extracurriculars (e.g., “Are you back to school like how you were before the injury?”). As there are currently no clinical measures or biomarkers to determine if someone has recovered from their concussion, the Recovery Screen will be the primary measure that will determine if participants are in the PSaC group or recovered control group. The Recovery Screen is completed by participants when the follow-up version of the HBI is completed, as well as at the imaging appointment at SickKids.

#### Neuropsychological Assessment

A neuropsychological assessment (Table 1) and self-report questionnaires (Table 2) will be administered to characterize participant cognitive, emotional, and behavioural profiles. Caregivers will complete parent-report versions of questionnaires (Table 3). The total duration for completion is 3-4 hours for youth and 40 minutes for caregivers. For caregivers who are unable to attend the appointment with youth, questionnaires are given in a mail-back envelope to be completed by the caregiver at home.

**Table 1.**
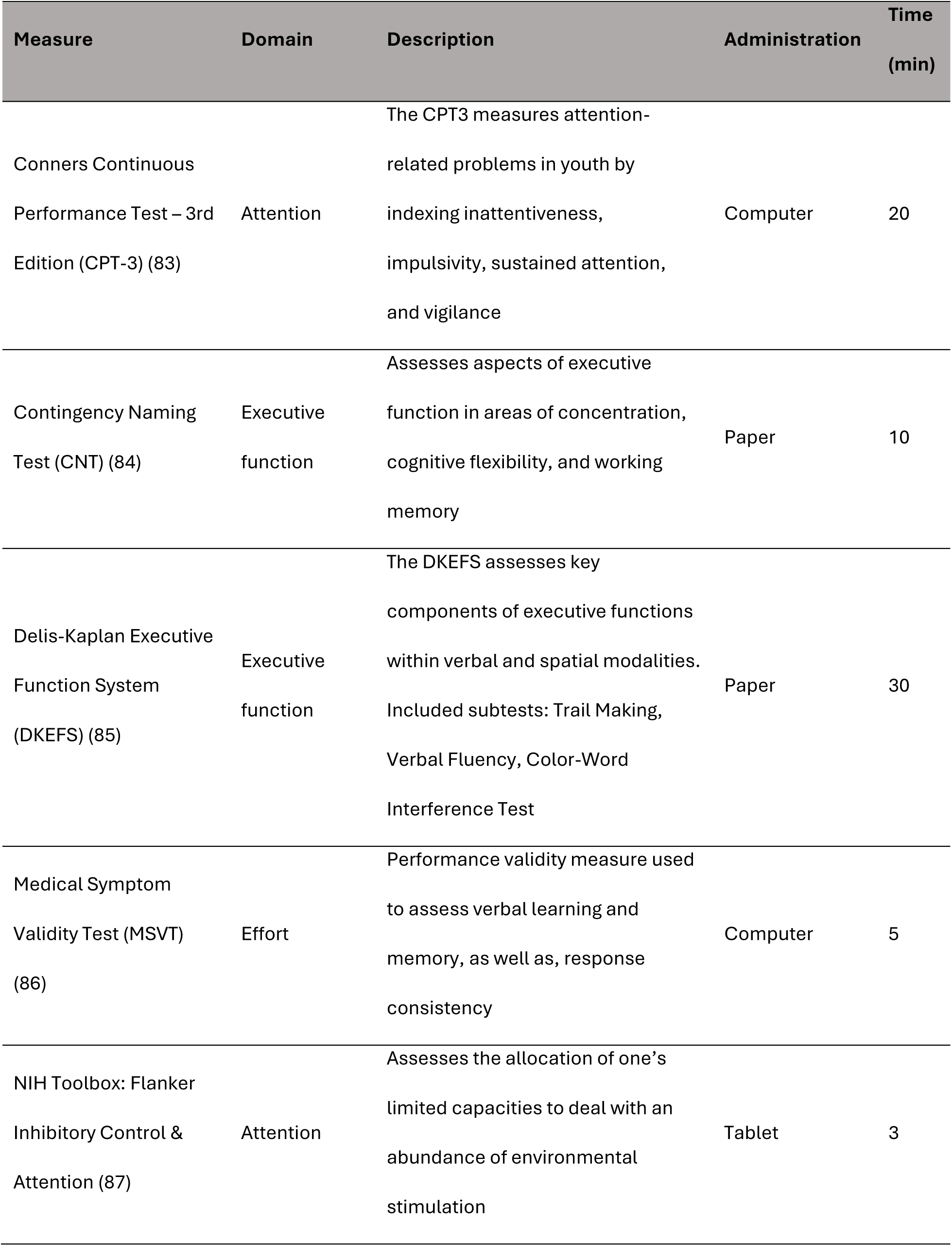
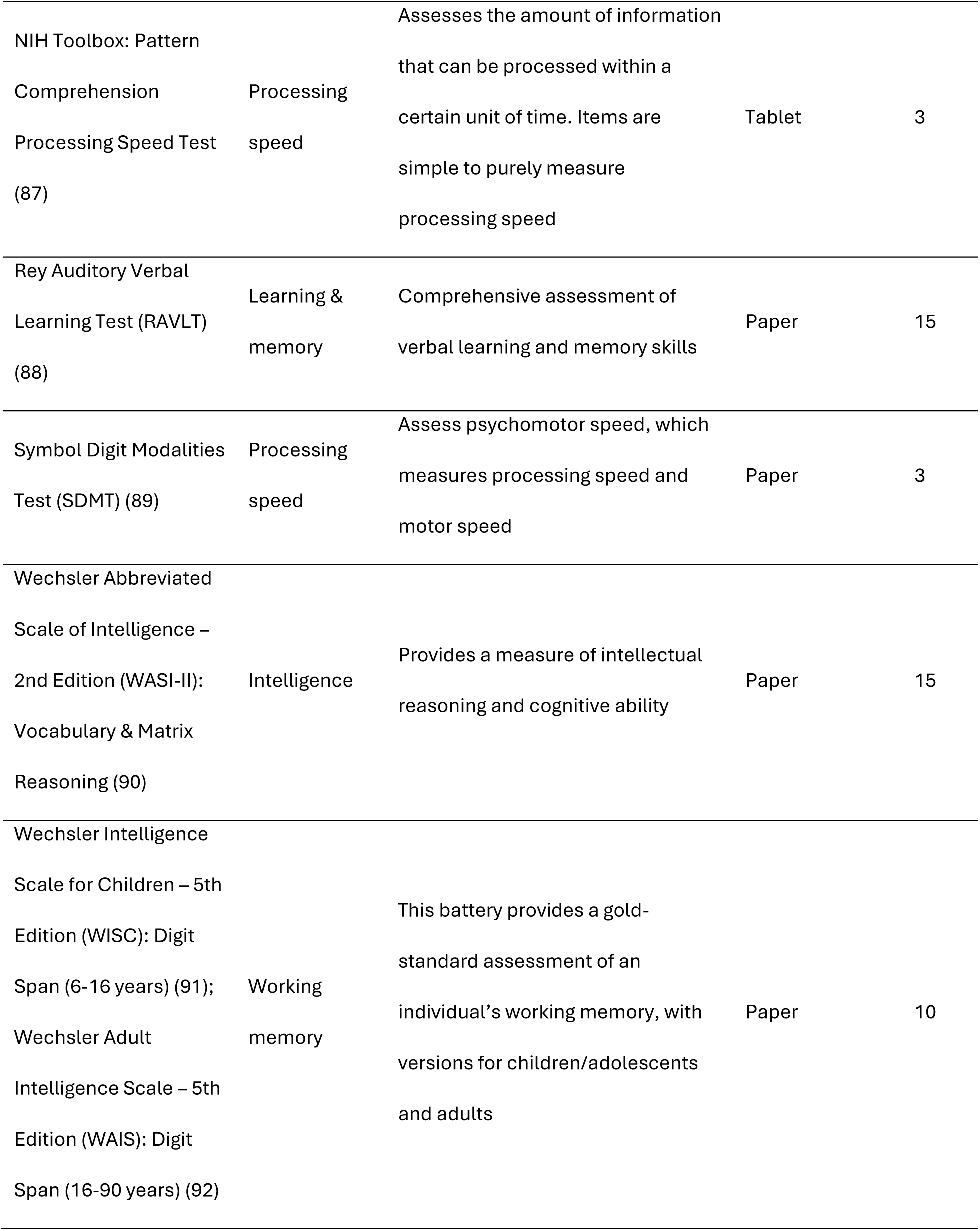
Neuropsychological assessment administered to youth participants.

| Measure | Domain | Description | Administration | Time (min) |
| --- | --- | --- | --- | --- |
| Conners Continuous Performance Test – 3rd Edition (CPT-3) (83) | Attention | The CPT3 measures attention-related problems in youth by indexing inattentiveness, impulsivity, sustained attention, and vigilance | Computer | 20 |
| Contingency Naming Test (CNT) (84) | Executive function | Assesses aspects of executive function in areas of concentration, cognitive flexibility, and working memory | Paper | 10 |
| Delis-Kaplan Executive Function System (DKEFS) (85) | Executive function | The DKEFS assesses key components of executive functions within verbal and spatial modalities. Included subtests: Trail Making, Verbal Fluency, Color-Word Interference Test | Paper | 30 |
| Medical Symptom Validity Test (MSVT) (86) | Effort | Performance validity measure used to assess verbal learning and memory, as well as, response consistency | Computer | 5 |
| NIH Toolbox: Flanker Inhibitory Control & Attention (87) | Attention | Assesses the allocation of one's limited capacities to deal with an abundance of environmental stimulation | Tablet | 3 |
| NIH Toolbox: Pattern Comprehension Processing Speed Test (87) | Processing speed | Assesses the amount of information that can be processed within a certain unit of time. Items are simple to purely measure processing speed | Tablet | 3 |
| Rey Auditory Verbal Learning Test (RAVLT) (88) | Learning & memory | Comprehensive assessment of verbal learning and memory skills | Paper | 15 |
| Symbol Digit Modalities Test (SDMT) (89) | Processing speed | Assess psychomotor speed, which measures processing speed and motor speed | Paper | 3 |
| Wechsler Abbreviated Scale of Intelligence – 2nd Edition (WASI-II): Vocabulary & Matrix Reasoning (90) | Intelligence | Provides a measure of intellectual reasoning and cognitive ability | Paper | 15 |
| Wechsler Intelligence Scale for Children – 5th Edition (WISC): Digit Span (6-16 years) (91); Wechsler Adult Intelligence Scale – 5th Edition (WAIS): Digit Span (16-90 years) (92) | Working memory | This battery provides a gold-standard assessment of an individual's working memory, with versions for children/adolescents and adults | Paper | 10 |

**Table 2.** Questionnaires administered to youth participants.

| Measure | Domain | Description | Administration | Time (min) |
| --- | --- | --- | --- | --- |
| Behaviour Rating Intervention of Executive Function – 2<sup>nd</sup> Edition (BRIEF-2) (93) | Behaviour & executive function | Self-report questionnaire that assesses behaviour and executive function in daily life with three core regulation indices (Behavior, Emotion, Cognitive). | Paper | 15 |
| Child-Adolescent Perfectionism Scale (CAPS) (94) | Perfectionism | A self-report measure of interpersonal expression of perfectionistic behavior for children and adolescents. | REDCap | 5 |
| Child Depression Inventory 2 (CDI-2) (95) | Emotions & Behaviour | Self-report form that helps assess cognitive, affective, and behavioural signs of depression in youth | Paper | 15 |
| Multidimensional Anxiety Scale for Children – 2<sup>nd</sup> Edition (MASC-2) (96) | Emotions & Behaviour | Self-report form to assess the presence of symptoms related to anxiety disorders in youth | Paper | 10 |
| Youth Demographic Questionnaire | Demographic & History | Structured to probe the adolescent's general medication history, current concussion symptoms, and developmental history | REDCap | 5 |
| Youth Self-Report (YSR) (97) | Emotions & Behaviour | Self-report form to assess behavioural and emotional problems in youth | Paper | 15 |

**Table 3.**
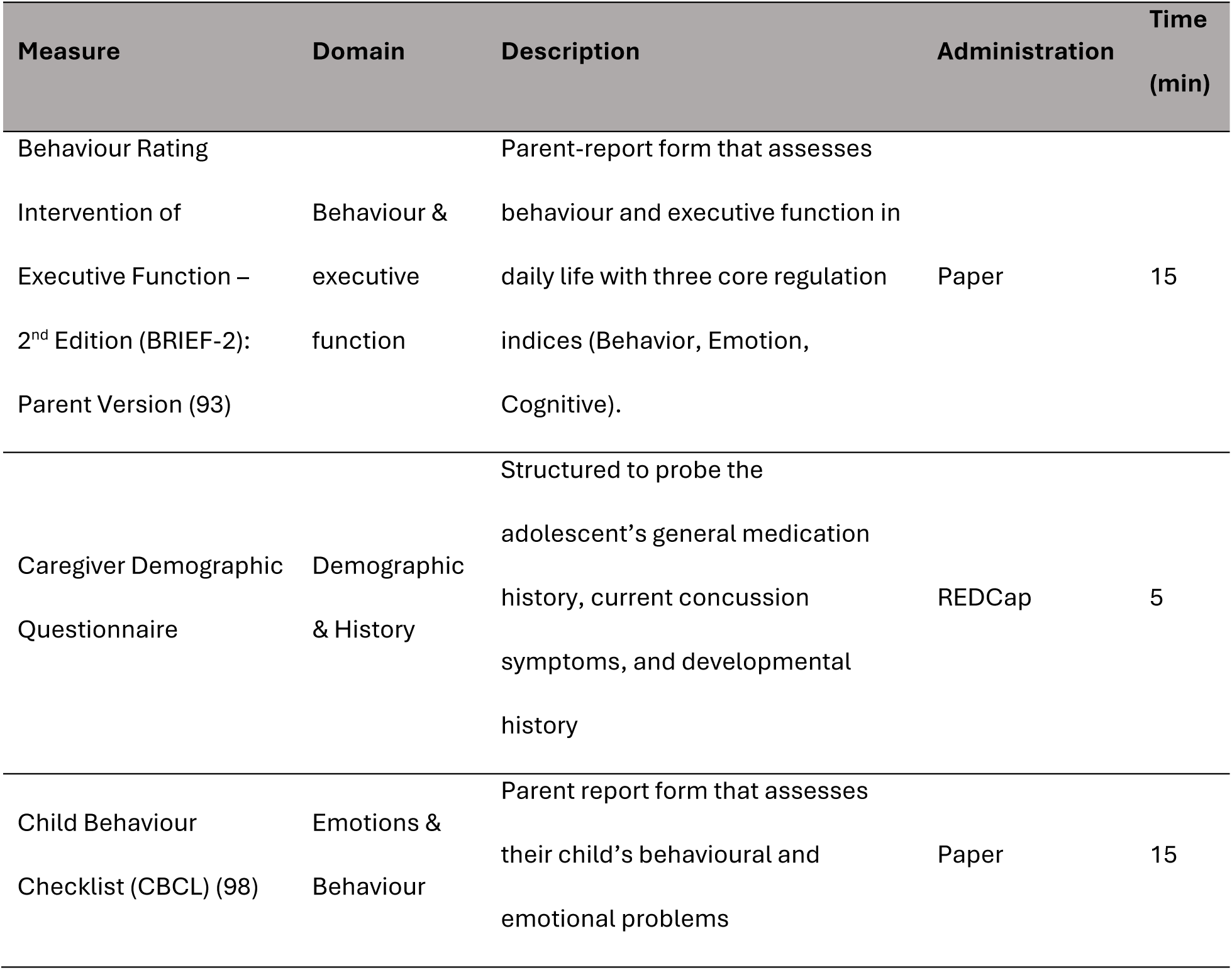
Questionnaires administered to caregiver participants.

| Measure | Domain | Description | Administration | Time (min) |
| --- | --- | --- | --- | --- |
| Behaviour Rating Intervention of Executive Function – 2<sup>nd</sup> Edition (BRIEF-2): Parent Version (93) | Behaviour & executive function | Parent-report form that assesses behaviour and executive function in daily life with three core regulation indices (Behavior, Emotion, Cognitive). | Paper | 15 |
| Caregiver Demographic Questionnaire | Demographic & History | Structured to probe the adolescent's general medication history, current concussion symptoms, and developmental history | REDCap | 5 |
| Child Behaviour Checklist (CBCL) (98) | Emotions & Behaviour | Parent report form that assesses their child's behavioural and emotional problems | Paper | 15 |

#### MRI protocol

All MRI sequencing will be carried out on a 3T Siemens Magnetom Prisma magnet located in the Research MRI Facility at SickKids. Overall, imaging of brain structure will provide information reflecting global and regional brain volumes, as well as tissue microstructure and tissue properties indicative of biological processes, such as myelination and cerebrospinal fluid flow. The total MRI scan time will be 1 hour, with scans and set-up (Table 4).

**Table 4.**
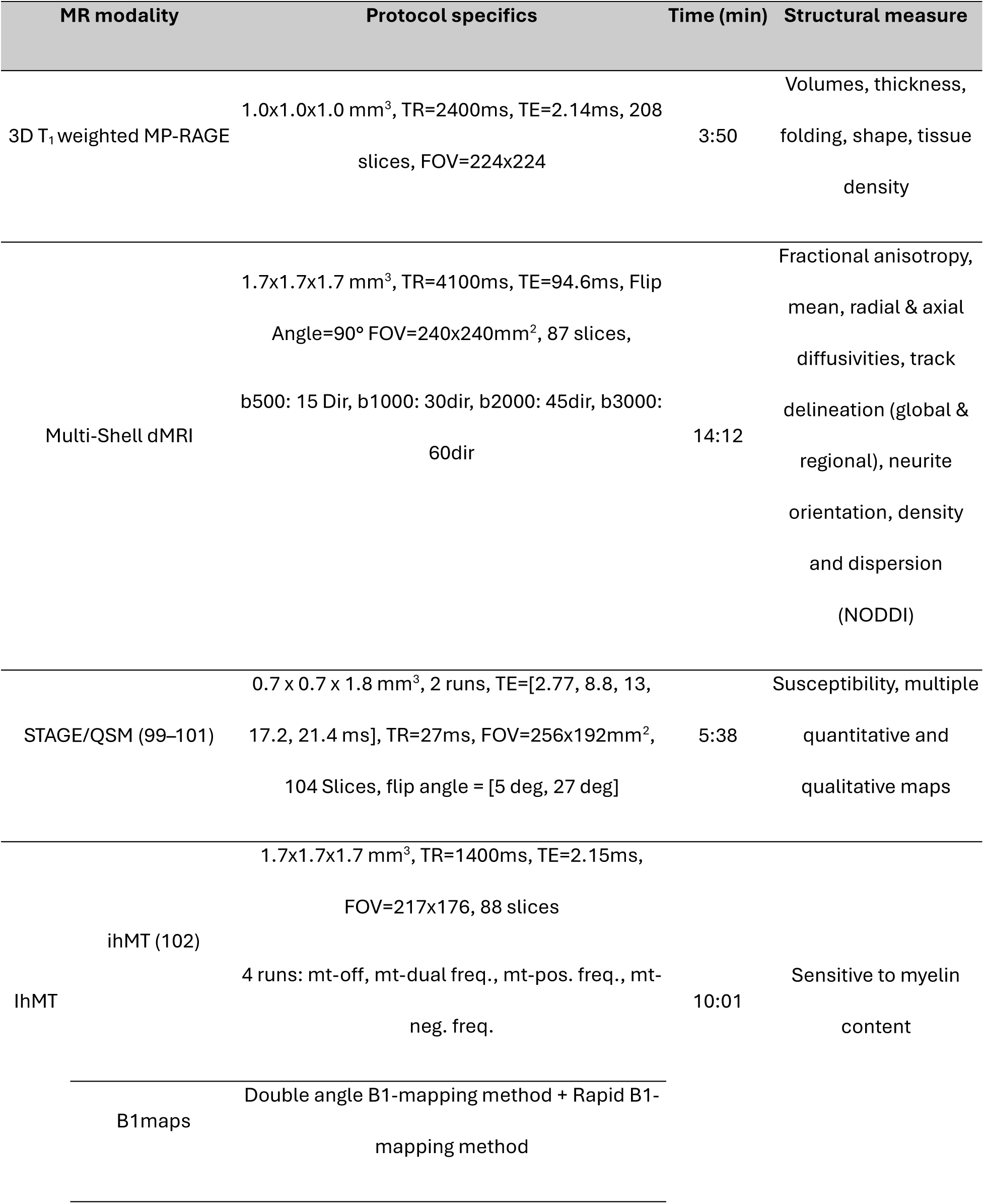
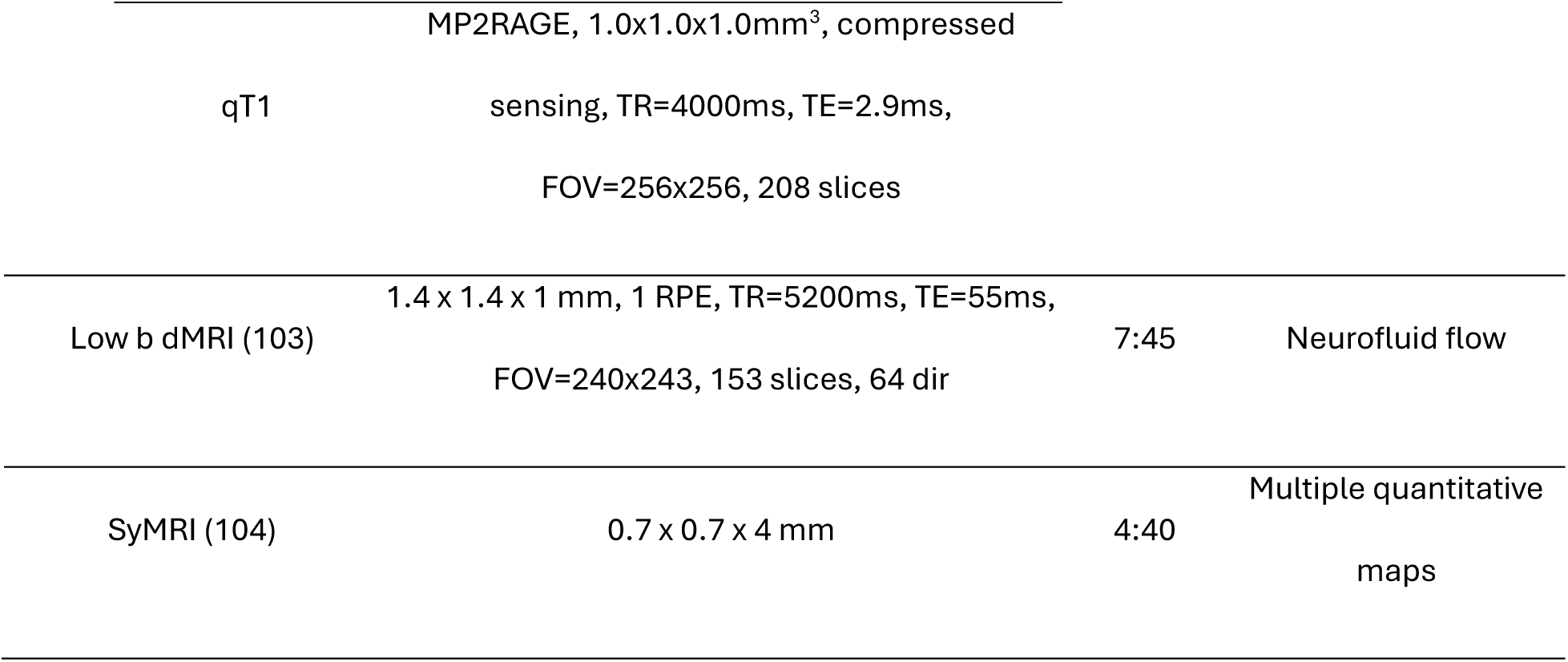
MRI Protocol.

#### MEG protocol

The MEG data will be collected with a CTF 151 channel system (CTF MSL, Coquitlam, BC, Canada) at SickKids during resting state and two experimental tasks. When the participant enters the MEG suite, they will be shown the system and the experimental procedures will be explained simply. Inside the magnetically shielded room (MSR), the participant will lie supine on a bed and place their head in the MEG helmet containing gradiometers. The helmet is large enough to accommodate almost all head sizes. The MEG data will be continuously recorded at a sampling rate of 600 Hz. The total MEG scan time is 30 minutes (scans + set-up).

##### Scan 1 – Resting state (5 minutes eyes open)

Participants will fixate on a central crosshair and stay “within their own thoughts” for the scan period, in the absence of any particular task.

##### Scan 2 – Sustained attention visuomotor task (8 minutes)

The stimuli will consist of a moving set of high-contrast (black and white) concentric circles. Participants will be instructed to maintain fixation at the centre of the circles, and to maintain attention while pressing a response key as fast and accurately as possible at the termination of each stimulation period (e.g., moving circles disappear). The stimulus will be presented for 1.5–2s followed by 2s of fixation cross only. Eighty stimuli will be presented in a session.

##### Scan 3 – Mental ffexibility task (c-8 minutes)

Using a 2-alternative forced choice design, the Intra-Extra Dimensional Set-Shifting task requires participants to match a designated target stimulus (a simple coloured geometric shape) to one of two other stimuli (a matching ‘prime’ target and a non-matching ‘prime’ distractor) on one of two dimensions, either colour or shape. After a randomised number of trials (3–5), the designated target stimulus will change and matching continues either in the same dimension (intradimensional shift; e.g. colour-to-colour) or in the other dimension (extradimensional e.g. colour-to-shape, or shape-to-colour). One hundred stimuli will be presented.

### Data Analysis

#### Objective 1

1) Identify functional and microstructural properties in intrinsic connectivity networks, as well as neuropsychological outcomes, that differentiate adolescents with concussion with PSaC from recovered controls.

Functional and microstructural measures will be compared between PSaC and recovered control groups using multivariate linear models. Primary functional imaging measures from MEG are oscillatory power and phase synchrony calculated on a voxel wise basis within (for power) and between regions (for phase synchrony, using e.g. weighted phase lag index wPLI). Primary structural imaging measures from diffusion MRI are the orientation dispersion index (ODI) and restricted diffusion calculated on a voxel-wise basis at 2mm beneath the cortex to capture superficial white matter. All images will be registered to a common Schaefer parcellation atlas (105) that delineates intrinsic connectivity networks. Mean measures will be computed within each intrinsic connectivity network and compared between groups to determine the most impacted by PSaC. The primary neuropsychological outcomes of interest are attention measures captured via the CPT-3. A similar comparison approach between groups will follow. For all comparisons, covariates will include sex, age, SES and others established after analysis of demographic and historical information. Multiple comparisons will be accounted for by applying a 5% false discovery rate correction (106).

#### Objective 2

2) Determine whether neural activity within intrinsic connectivity networks mediates the relationship between network microstructure and distinct symptoms and neuropsychological outcomes in adolescents with concussion.

Mediation models will be set up to test if functional measures mediate the relationship between microstructural measures and neuropsychological outcomes within intrinsic connectivity networks. Mediation models will be built using the following specifications: independent variable = microstructural measure (e.g. ODI or restricted diffusion); dependent variable = neuropsychological symptom (e.g. sustained and executive attention, anxiety); mediator = neural activity (oscillatory power, wPLI); covariates = sex, age, SES and other covariates established after analysis of demographic and historical information. Models will be built using mean functional and microstructural measures in intrinsic connectivity networks (e.g. frontoparietal network, salience network). The effect size (ab) of the mediator will be calculated from the product of the partial correlations of each coefficient (a and b). The Sobel test will be conducted to test the significance of this effect.

### Statistical Power

The sample size is determined based on the primary group-comparison analyses in Objective 1. With a total planned enrollment of N = 160 (PSaC = 80; recovered controls = 80), the study will have 80% power to detect a small-to-moderate effect size (Cohen’s d = 0.45) at a two-sided significance level of 0.05. This effect size is consistent with prior diffusion MRI and MEG studies of concussion (59,107,108). Mediation analyses in Objective 2 will be conducted as secondary mechanistic analyses with the full sample of 160 participants, recognizing that indirect effects are typically smaller.

## Discussion

### Expected Impact

Given the high prevalence of concussion and long-lasting impairments experienced by adolescents with PSaC, understanding how disrupted functional and structural brain circuits underpin PSaC will provide essential guidance for precision treatment strategies for youth with PSaC, including targeted brain-based treatments (61,109–118). Brain stimulation therapies have proven to be effective in potentiating plasticity and repair in complex neurological and psychiatric conditions (e.g., stroke, depression) (114–116,118); with some beneficial effects on cognitive functioning and attention (113). Our approach will identify microstructural and functional deficits within relevant networks to accurately target therapeutic neurostimulation protocols.

### Dissemination plans

A multi-pronged mobilization strategy that consists of integrated and end-of-grant approaches will be used to widely disseminate research findings with intended audiences, including clinicians, researchers, and youth with PSaC and their families. The results of this study will be published in relevant peer-reviewed scientific journals. The study findings will be presented at national and international scientific conferences in the field of brain injury. Results will be shared with clinical brain injury teams at HBKRH and SickKids. A lay language summary of the study results will be presented to participants at the end of the project.

### Strengths and limitations

Strengths of this study include a novel multimodal study design integrating MEG, MRI, and neuropsychological assessment. Further, advanced analytical techniques, such as neurite orientation dispersion and density imaging (NODDI), will provide better explanations of observed white matter abnormalities, as they have been shown to be sensitive to persistent axon damage and inflammation (119).

It is important to acknowledge that this clinical population is highly heterogenous, in terms of symptoms, recovery experiences, and adolescent development. To account for this and potential confounders, we will collect demographic and history data through detailed questionnaires. Some variables will include concussion history, mechanism of injury, concussion symptoms, comorbid conditions (e.g., anxiety, depression, psychiatric), and developmental considerations. We will collect sex at birth and have participants self-report gender. Further, the sample may not represent a community sample, as the primary recruitment mechanism is hospital settings. This may result in the enrollment of individuals who are more severely impacted by their concussion.

### Data management

All data will be stored using institutionally approved methods at HBKRH and SickKids. A Data Transfer Agreement (DTA) has been executed to facilitate the transfer of raw neuropsychological and neuroimaging data between study sites.

Data from the neuropsychological assessment and questionnaires will be stored in both hard and electronic formats, as some measures are administered on paper while others are administered on a computer. Hard copies of participant files will be stored in a locked cabinet at HBKRH. Electronic copies of measures (MSVT, CPT-3, Flanker Inhibitory Control & Attention, Pattern Comprehension Processing Speed Test) will be stored in a controlled-access folder on HBKRH’s network with firewalls and security in place. All measures will be input into a REDCap database, and data will be downloaded and stored on the HBKRH hospital network.

Imaging data (MRI and MEG) collected at SickKids will be stored on the Research IT High Performance Computing Service (RIT-HPC). The RIT-HPC is a core facility at SickKids that provides infrastructure to run advanced applications efficiently, reliably, and quickly, as well as to store files securely.

### Ethical Considerations

This study received ethical approval at both sites through Clinical Trials Ontario (CTO#4351), with the HBKRH Research Ethics Board as the REB of record. All participants will provide informed written consent/assent before enrolling in the study.

### Safety Considerations

Participants will be completing questionnaires pertaining to mental health and therefore may report a history or current thoughts of self-harm. If a participant expresses serious mental health concerns or endorses these thoughts or behaviours on questionnaires, the research team member conducting the data collection will follow the established Risk and Distress protocol. Briefly, the participant will be directly approached by a clinically trained member of the research team to discuss the reported concerns prior to them leaving the data collection session. They will be encouraged to discuss their mental health concerns with their family physician or other mental health professionals and will be provided with a list of community mental health resources. If the participant endorses immediate concerns of self-harm or suicide risk, the clinically trained research team member will assess the situation. If the participant has a suicide risk plan in place, the clinician will defer to that plan. If the participant does not have a risk plan in place, the clinician will trust the caregiver to escort the youth to the nearest emergency department. Alternatively, the clinician will call emergency services to escort the participant to the nearest emergency department.

Additionally, as with all MRI studies, there is risk of incidental and/or actionable findings after completion of the scan. Any material incidental findings will be outlined by a radiologist in a full report, which will be communicated with the participant through one of the following pathways: 1) if the participant is a client at HBKRH or SickKids, the co-principal investigators will discuss the delegation of responsibility for disclosure, 2) if the participant is not a client at HBKRH or SickKids, a copy of the radiology report and imaging will be sent to their family physician, or 3) if the participant does not have a family physician, a copy of the radiology report will be provided to the contact person identified by the participant and the participating caregiver. If using the third pathway, the research team will recommend that the participant seek care from a walk-in clinic to discuss the radiology report with a physician and to help facilitate appropriate referrals.

### Study Status

This study was awarded funding on October 1^st^, 2022 (CIHR #183821). Ethical approval was obtained February 7^th^ 2024 and recruitment began April 1^st^, 2024. The estimated end date of recruitment is March 31^st^, 2027. The study was registered with OSF on August 24^th^, 2026 (Registration DOI: https://doi.org/10.17605/OSF.IO/RKUM6).

## Data Availability

No datasets were reported or analysed during the current study. The data availability policy is not applicable.

## Authors’ contributions

## Conceptualization

Shannon E. Scratch, Benjamin Dunkley, Anne Wheeler.

## Data curation

Pavreet Gill, Alexa Irvin, Rouzbeh Zamyadi.

## Funding acquisition

Shannon E. Scratch, Adrienne L. Davis, George Ibrahim, Benjamin Dunkley, Anne Wheeler.

## Investigation

Shannon E. Scratch, Pavreet Gill, Andrea Hickling, Alexa Irvin, Benjamin Dunkley, Anne Wheeler.

## Methodology

Shannon E. Scratch, Pavreet Gill, Andrea Hickling, Alexa Irvin, Shireen Parimoo, Rouzbeh Zamyadi, Phillip R. Johnston, Benjamin Dunkley, Anne Wheeler.

## Project administration

Pavreet Gill, Andrea Hickling, Alexa Irvin.

## Resources

Shannon E. Scratch, Benjamin Dunkley, Anne Wheeler.

## Supervision

Shannon E. Scratch, Benjamin Dunkley, Anne Wheeler.

## Visualization

Shannon E. Scratch, Pavreet Gill, Andrea Hickling, Alexa Irvin, Benjamin Dunkley, Anne Wheeler.

## Writing – original draft

Shannon E. Scratch, Pavreet Gill, Andrea Hickling, Alexa Irvin, Benjamin Dunkley, Anne Wheeler.

## Writing – review s editing

Shannon E. Scratch, Pavreet Gill, Andrea Hickling, Alexa Irvin, Adrienne L. Davis, George Ibrahim, Shireen Parimoo, Rouzbeh Zamyadi, Phillip R. Johnston, Benjamin Dunkley, Anne Wheeler.

## Acknowledgements

We would like to acknowledge the efforts of the members of the NOvEL Lab Team (Bloorview Research Institute), specifically Kylie Mallory, Hussain Syed and Elena Sheldrake. We would also like to acknowledge the efforts of the members of the BRANCH Lab and Dunkley Lab (SickKids Research Institute). We would also like to thank the SickKids Emergency Assistants for Research in Child Health (SEARCH) team for their support with recruitment for this study.

## Data Availability

No datasets were generated or analysed during the current study. All relevant data from this study will be made available upon study completion.

